# Association between PM2.5 air pollution, temperature, and sunlight during different infectious stages with the case fatality of COVID-19 in the United Kingdom: a modeling study

**DOI:** 10.1101/2023.04.07.23288300

**Authors:** M. Pear Hossain, Wen Zhou, Marco Y. T. Leung, Hsiang-Yu Yuan

## Abstract

Although the relationship between the environmental factors such as weather conditions and air pollution and COVID-19 case fatality rate (CFR) has been found, the impacts of these factors to which infected cases are exposed at different infectious stages (e.g., virus exposure time, incubation period, and at or after symptom onset) are still unknown. Understanding this link can help reduce mortality rates. During the first wave of COVID-19 in the United Kingdom (UK), the CFR varied widely between and among the four countries of the UK, allowing such differential impacts to be assessed.

We developed a generalized linear mixed-effect model combined with distributed lag nonlinear models to estimate the odds ratio of the weather factors (i.e., temperature, sunlight, relative humidity, and rainfall) and air pollution (i.e., ozone, *NO*_2_, *SO*_2_, *CO, PM*_10_ and *PM*_2.5_) using data between March 26, 2020 and May 12, 2020 in the UK. After retrospectively time adjusted CFR was estimated using back-projection technique, the stepwise model selection method was used to choose the best model based on Akaike information criteria (AIC) and the closeness between the predicted and observed values of CFR.

We found that the low temperature (8-11°C), prolonged sunlight duration (11-13hours) and increased *PM*_2.5_ (11-18 *μg*/*m*^3^) after the incubation period posed a greater risk of death (measured by odds ratio (OR)) than the earlier infectious stages. The risk reached its maximum level when the low temperature occurred one day after (OR = 1.76; 95% CI: 1.10-2.81), prolonged sunlight duration 2-3 days after (OR = 1.50; 95% CI: 1.03-2.18) and increased *P*.*M*_2.5_ at the onset of symptom (OR =1.72; 95% CI: 1.30-2.26). In contrast, prolonged sunlight duration showed a protective effect during the incubation period or earlier.

After reopening, many COVID-19 cases will be identified after their symptoms appear. The findings highlight the importance of designing different preventive measures against severe illness or death considering the time before and after symptom onset.

## Introduction

The emergence of COVID-19 has led to an unprecedented number of infections and deaths worldwide. Certain environmental factors, such as weather conditions and air pollution, have been shown to influence disease severity. Knowing the consequence of these factors to which infected individuals are exposed at different infectious stages (e.g., virus exposure time, incubation period, and at or after symptom onset) can potentially help to form guidance on reducing the number of COVID-19 deaths. Unfortunately, the evidence of such differential effects on case fatality rate (i.e., the probability of death after infection) remains largely unknown.

Recent population studies have reported the association between COVID-19 deaths and weather conditions, such as temperature and humidity (Benedetti et al. 2020; Ma et al. 2020; Wu et al. 2020b). A colder condition can increase the viability and survival of viruses during disease transmission (for review, see (Mecenas et al. 2020)), leading to a higher viral load. The viral load has been demonstrated to be associated with disease severity (Fajnzylber et al. 2020). Another possible route to affect disease mortality by temperature and humidity is through modulating immune responses. Studies have found that overreaction of immune responses, such as cytokine storm, triggered by innate immunity, can lead to severe consequences after the infection. Furthermore, the activity of macrophages, which drives innate immunity, has been shown to be associated with temperature (Hardie et al. 1994; Hassan et al. 2020). This innate defense mechanism generally began after the incubation period (Schultze and Aschenbrenner 2021). Hence, exposure to environmental factors at or after symptom onset might contribute to such dysregulated innate immunity.

In addition to temperature, sunlight exposure is another potential environmental risk factor for COVID-19 deaths. Lower vitamin D levels were associated with an increased risk of infection and its severity (Merzon et al. 2020; Panagiotou et al. 2020). Sunlight exposure aids in synthesizing vitamin D, which is likely to reduce the severity of COVID-19 (Laird et al. 2020; Martineau and Forouhi 2020). Presumably, the effect of sunlight has to occur in the early infectious stages in order to influence immune response. However, no studies have shown at which infectious stages, sunlight exposure is associated with COVID-19 mortality.

Exposure to ambient air pollution is also associated with the transmissibility, population susceptibility, and severity of COVID-19 (Liang et al. 2020; Stieb et al. 2021; Woodby et al. 2021). The main components of air pollution are gases and particles such as carbon monoxide (CO), nitrogen dioxide (*NO*_2_), sulphur dioxide (*SO*_2_), ozone (*O*_3_), and particulate matter of size ≤ 10 *μm* (*PM*_10_) and ≤ *μm* (*PM*_2.5_), respectively. As a result, air pollution is considered as the transport of viral particles in the air (Frontera et al. 2020; Martelletti and Martelletti 2020) and within the respiratory tract. By worsening chronic respiratory diseases or modulating immune responses, air pollution could increase the severity of COVID-19 (Bourdrel et al. 2021). Therefore, it is important to understand the impact of air pollution on disease fatality when they are exposed after the incubation period.

Worldwide, the United Kingdom (UK) has the second highest number of COVID-19 related deaths as of May 31, 2020 (37,175 deaths) (Our World in Data 2021). After the initial spread of COVID-19, a strict social distancing policy was implemented on March 26 in the UK constituent countries, except Northern Ireland which two days later also adopted the same policy, to reduce COVID-19 transmission. This lockdown was initially relaxed on May 13, 2020. Despite the similar lockdowns, by the end of May, 2020, of its four constituent countries (England, Northern Ireland, Scotland and Wales), the most significantly affected country was England.

This study aimed to identify risk factors among weather conditions and air pollution and quantify the impacts of these factors at different stages of infections on the probability of death after COVID-19 infection during the early spread in the UK. We developed a generalized linear mixed-effect model with distributed lag nonlinear models (DLNM) to assess the risk of environmental factors at the different stages. The results may help to inform recommendations of preventive measures for reducing disease severity.

## Material and Methods

### Epidemiological data

We collected the daily reported cases and deaths from a publicly available source (GOV.UK 2021). To assess the impact of environmental factors on the severity of COVID-19, the study period was defined as the time between March 26, 2020, and May 12, 2020, when the intensity of non-pharmaceutical interventions was relatively stable and similar between different countries in the UK (i.e., during the first lockdown period) (The Institute for Government 2020). Therefore, the number of deaths was not largely affected by the changes in control measures.

### Environmental data

Weather data were collected from the European Climate Assessment and Dataset (ECA&D) project (Tank et al. 2002). The daily mean temperature was obtained from 120 UK meteorological stations, while mean sunlight duration was available from 24 stations. The temperature data had 1.1% values missing. To address these missing observations, we calculated the average of the temperatures of the previous 7 days to replace the missing values. As relative humidity data were not directly available from ECA&D at the time of data collection, we collected dew point temperatures from the National Oceanic and Atmospheric Administration (NOAA) (National Centers for Environmental Information 2020) to calculate relative humidity following a previous method (McNoldy 2020). Air pollution data, such as *CO, NO*_2_, *SO*_2_, *O*_3_, *PM*_10_ and *PM*_2.5_, were collected from Air Information Resource in the Department for Environment Food and Rural Affairs, UK (Department for Environment Food and Rural Affairs 2021).

### Back-projection of COVID-19 deaths and estimation of instantaneous CFR

In order to estimate the probability of newly confirmed infected cases who die later due to the infections on a given day, instantaneous case fatality rate (iCFR) was used (Liang and Yuan 2022). One way to calculate iCFR is through a non-parametric back-projection approach to retrospectively adjust the time of death cases (Becker et al. 1991). This reduces the possible bias caused by different time points between reporting of cases and deaths when calculating the rate.

We assumed that COVID-19 transmission dynamics appeared in different disease status including as exposed (*E*), symptom onset (*I*), cases confirmation (*C*) and deaths (*D*) (Figure 1A). We assumed the time span between exposure and symptom onset to be days (referred as incubation period), and the time between symptom onset and case confirmation to be *t*_1_ = 5.71 days (referred as confirmation delay). Additionally, the duration between case confirmation and death (time to death) was taken to be *t*_3_ = 7.92 days. These values were estimated in our recent study (Liang and Yuan 2022). Given the time to death follows a gamma distribution, with a mean of *t*_3_ = 7.92 days, we retrospectively calculated the actual number of deaths, (*D*^′^), which were likely to be members of confirmed cases using an R function backprojNP (Meyer et al. 2017). Finally, iCFR was calculated as a ratio of *D*^′^ and *C*.

**Figure 1.**
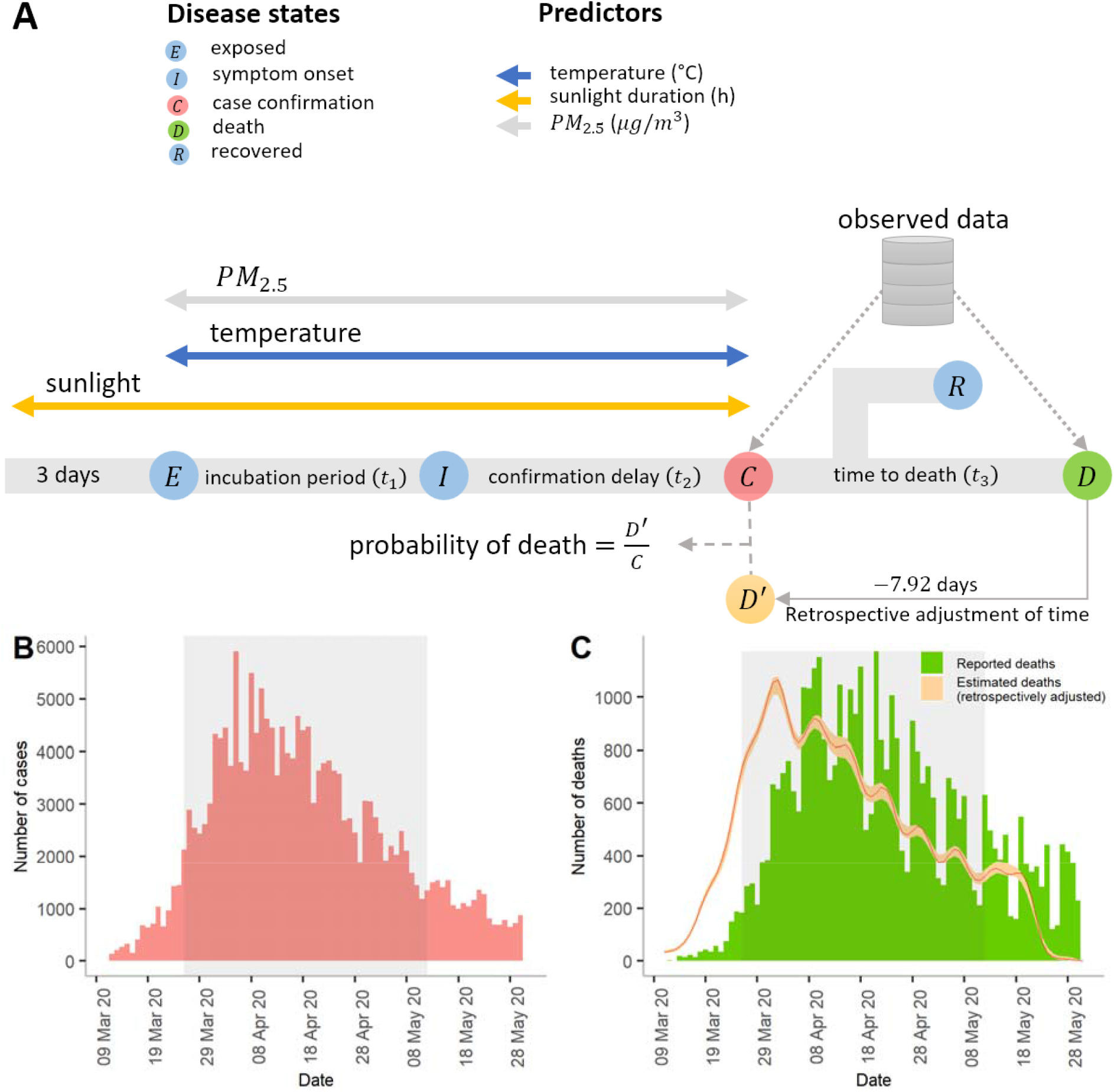
(A) Timeline of disease states and environmental risk factors. ,,, and represent the disease states such as exposed, symptom onset (or the end of incubation period), case confirmation, recovered and reporte deaths, respectively. Number of deaths were assumed a subset of infected cases who were reported previous days. Hence, a retrospective adjustment of time was made for estimating deaths who were reported as positive cases at time using non-parametric back-projection method. These estimated deaths were labeled as, and therefore for each day. Thus, the iCFR is estimated as the ratio of and. **(B) and (C) reported cases and deaths in the UK**. Bar charts representing the daily confirmed cases and deaths, respectively. The red line in (C) represents the retrospectively estimated number of confirmed cases who later died per day and the yellow area represents the corresponding 95% confidence interval. The gray-shaded regions in (B) and (C) represent the duration of the lockdown in the UK.

### Model formulation

We used a generalized linear mixed-effect model (Gurka et al. 2012) with DLNM (Gasparrinia et al. 2010). We adjusted for the effects of relative humidity on the day of exposure to determine whether the iCFR was affected by it on the days when the indexed cases were exposed. We assumed the number of deaths, *D*^′^ follows a binomial distribution with a probability, 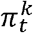 (the probability of death after infection), among confirmed cases *C*, i.e., 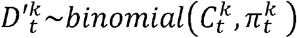, where *k* indicates a particular location and *t* represents a day. The model was developed as follows

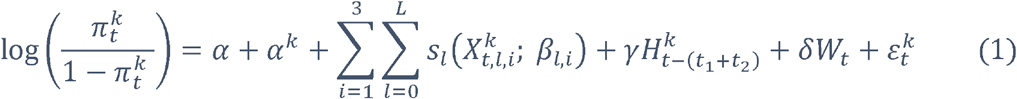

where 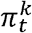 represents the expected iCFR among newly confirmed cases on day *t* at location *k* (*k* = 1,2,3 or 4; representing the four countries of the UK), *α* is the overall intercept of the model and *a*_*k*_ is the region-specific random intercept. *s*_*l*_ represents a smooth function of the environmental predictor 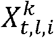 (*i*= 1,2,3; representing temperature, sunlight duration and *PM*_2.5_) and *l* represents the lag days from the day of confirmation to the day of exposure. *L* is the maximum lag, which was defined as the sum of the incubation period and confirmation delay, i.e., 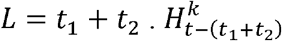 represents the relative humidity on the day of exposure at time *t* and location *k. W*_*t*_ represents the day of the week on a given day *t* which allows to adjust for weekly effect of COVID-19 testing whereby more test results are reported on specific days of the week (i.e., first day of the week or weekend). A random error term is represented by 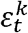. See detailed descriptions in Supplementary Information.

To completely capture the overall impact of weather during the incubation period and confirmation delay, we used a maximum lag of 10 days for temperature and air pollution. For sunlight duration, the time between three days before virus exposure and confirmation was considered under the assumption that vitamin D synthesis in individuals can happen before virus exposure and affect the immune response thereafter. The odds ratio of death was calculated using a reference value of each predictor. The linear effect of relative humidity was assessed on the day of exposure because models using distributed lagged effects of relative humidity did not show good fitting results based on Akaike information criterion (AIC).

### Model selection criteria

To identify the best model (best-prediction model) among different combinations of predictors, in a two-stage selection approach, AIC were used in the first stage to choose a set of candidate models using a stepwise selection approach (details in supplementary materials). The models that gave relatively lower AIC were considered the candidate models in the second stage. In the second stage, we compared the model’s output to the observed data. The model that produced the lowest root means square errors (RMSE) was chosen as the best model.

### Sensitivity analysis for model validation

The best model was further tested for sensitivity in terms of future prediction. We extended the data until mid-September 2020 when the first alpha variant was detected (Higgins-Dunn 2020). The data during the study period were trained in the model, and the data between May 13, 2020, and September 15, 2020, were considered test data sets. Finally, we estimated the prediction results of iCFR and compared them with the retrospectively time adjusted iCFR.

## Results

To estimate the iCFR, we first retrospectively adjusted the daily number of reported deaths to their possible confirmed data and divided this number by the daily number of confirmed cases. The reported deaths were back-projected to the time of confirmation assuming that infected individuals were died 7.92 days on average after they were confirmed (see Methods).

We observed variations in both the iCFR and environmental predictors, such as weather and air pollution, in the UK (Figure S1 and S2). The iCFR was highest at the beginning of the lockdown in each of the UK’s four countries, and the ratio gradually declined over time (Figure S1). Among them, England showed a highest iCFR. Temperature, sunlight duration, and humidity were low in England and Scotland at the start of the outbreak and declined later. Maximum fluctuation in the concentration of *PM*2.5 was found in England and Wales (Figure S2). The detailed description of the variation of these factors and CFR was described in Supplementary Information.

We compared seven models, from a baseline to more complicated models, including different combinations of the weather and air pollution predictors (Table 1). The best model (Model 6), including temperature, sunlight duration and *PM*_2.5_, was selected after showing that the AIC was low and the RMSE of the observed and the estimated values was the lowest than others (Table 2).

**Table 1.** Initial model selection. AIC and BIC represent Akaike information criterion and Bayesian information criteria, respectively. All candidate models were adjusted for the days of the week.

| No. | Model | AIC | BIC | Log-likelihood |
| --- | --- | --- | --- | --- |
| 1 | $\alpha + \alpha^k + \delta W_t$<br>Baseline (regional random effect) | 2967 | 2990 | -1475 |
| 2 | $\alpha + \alpha^k + \delta W_t + f(S)$<br>Regional random effect and sunlight duration | 650 | 708 | -301 |
| 3 | $\alpha + \alpha^k + \delta W_t + f(S) + f(T)$<br>Regional random effect, temperature, and sunlight duration | 595 | 690 | -258 |
| 4 | $\alpha + \alpha^k + \delta W_t + f(S) + f(T) + f(SO_2)$<br>Regional random effect, temperature, sunlight duration and sulfur dioxide | 581 | 705 | -238 |
| 5 | $\alpha + \alpha^k + \delta W_t + f(S) + f(T) + f(PM_{10})$<br>Regional random effect, temperature, sunlight duration and particular matter ($\leq 10 \mu m$) | 579 | 713 | -234 |
| 6 | $\alpha + \alpha^k + \delta W_t + f(S) + f(T) + f(PM_{2.5})$<br>Regional random effect, temperature, sunlight duration and particular matter ($\leq 2.5 \mu m$) | 578 | 712 | -233 |
| 7 | $\alpha + \alpha^k + \delta W_t + f(S) + f(T) + f(PM_{2.5}) + \gamma H_{t-(t_1+t_2)}^k$<br>Regional random effect, temperature, sunlight duration, particular matter ($\leq 2.5 \mu m$) and humidity | 579 | 715 | -232 |

**Table 2.** Model comparison. The three candidate models (Models 5-7) were compared using predicted results.

| Model | Model expression | RMSE |
| --- | --- | --- |
| Model 5 | $\alpha + \alpha^k + \delta W_t + f(S) + f(T) + f(PM_{10})$<br>Regional random effect, temperature, sunlight duration and particular matter ($\leq 10 \mu m$) | 0.02040 |
| Model 6 | $\alpha + \alpha^k + \delta W_t + f(S) + f(T) + f(PM_{2.5})$<br>Regional random effect, temperature, sunlight duration and particular matter ($\leq 2.5 \mu m$) | 0.01767 |
| Model 7 | $\alpha + \alpha^k + \delta W_t + f(S) + f(T) + f(PM_{2.5}) + \gamma H_{t-(t_1+t_2)}^k$<br>Regional random effect, , temperature, sunlight duration, particular matter ($\leq 2.5 \mu m$) and humidity | 0.01855 |

RMSE = Root mean squared error

The model successfully captured the pattern of iCFR in each country (Figure S3).

### Differential risks of environmental factors

We assessed the differential effects of temperature, sunlight duration and *PM*_2.5_ during the course of infection. Compared to the reference temperature of 12 °C, low temperatures between 8-11 °C after the incubation period were associated with a higher risk (measured by odds ratio) of death (Figure 2A). A temperature of 9.5 °C at 7 days after the exposed to virus gave a maximum OR of 1.76 (95% CI: 1.10-2.81). When temperatures were below 8 °C the death risk became lower both during and after the incubation period. Whether infected cases changed their behaviors, such as staying indoors more during those very cold days is unknown.

**Figure 2.**
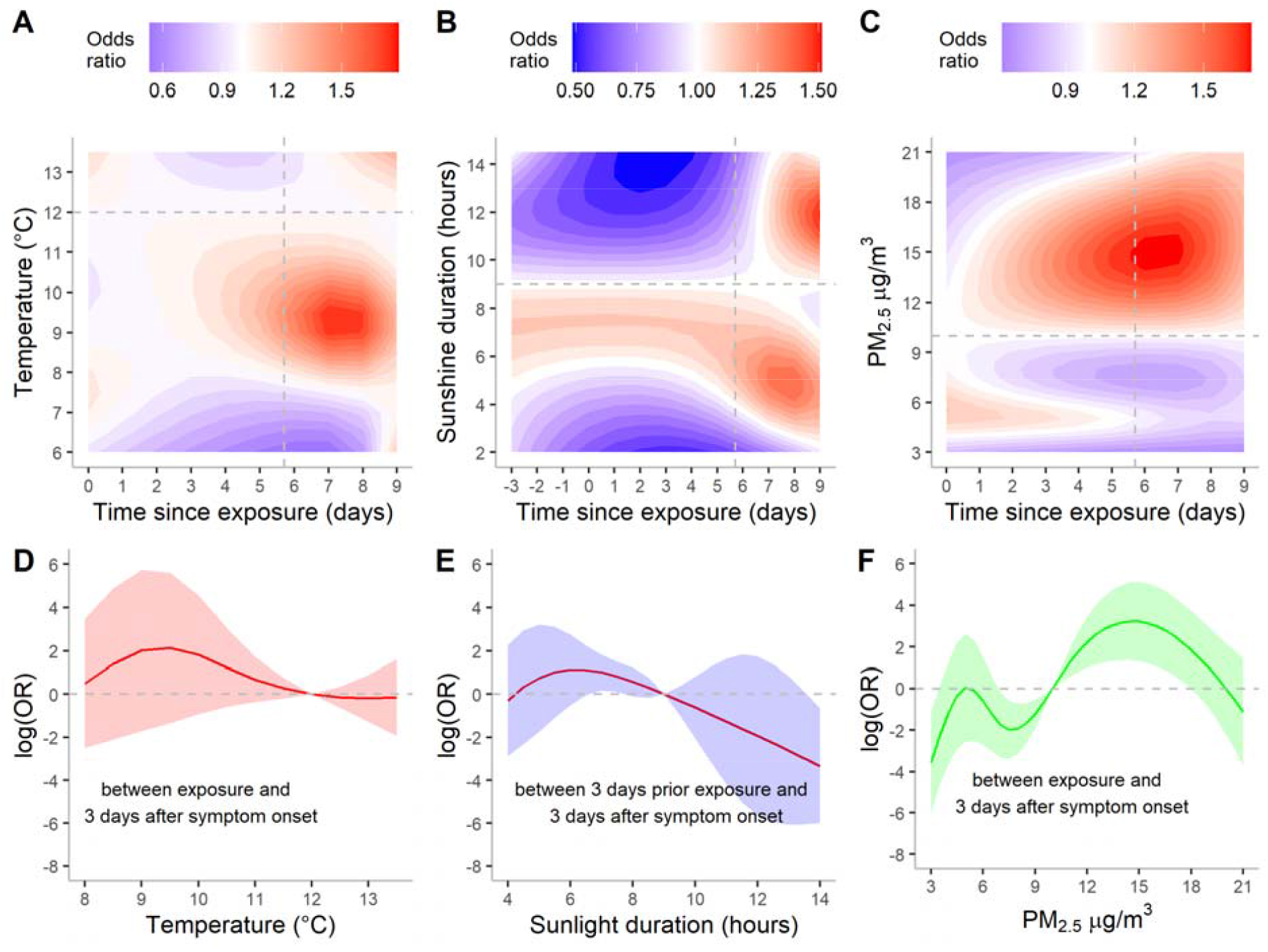
Risk of COVID-19 fatality under different environmental conditions and different time points since virus exposure (A, B, C). (A) Temperature, (B) sunlight duration and (C) particular matter (*PM*_2.5_). The vertical dashed lines in A, B, C represent the date of symptom onset (on day 5.71 since exposure). Therefore, time between 0 and 5.71 days represents the incubation period. Odds ratio was estimated with respect to the reference value (horizontal lines in A, B, C) of each predictor. Reference value for temperature was 12°C, sunlight duration 9h and *PM*_2.5_ 10 *μg*/*m*^3^. **Cumulative effects of environmental factors on the odds ratio of COVID-19 mortality (D, E, F)**. Horizontal dashed lines represent the baseline odds ratio at the reference values of the environmental predictors. The shaded regions represent 95% confidence interval of the log transformed odds ratio. The overall effect of temperature and *PM*_2.5_ was estimated for the duration between virus exposure and the confirmation day, whereas the cumulative effects of sunlight duration was estimated from three days prior to the virus exposure to the case confirmation day.

Furthermore, we found that the sunlight-fatality relationship was distinctly different before and after the estimated symptom onset (i.e., during and after the incubation period) (Figure 2B). The exposure to sunlight after the appearance of symptoms appeared to be more harmful. Prolonged sunlight exposure (11-13h) about 2 days after symptom onset was associated with a greater risk of death (OR = 1.50; 95% CI: 1.03-2.18). However, the prolonged exposure to sunlight, in contrast, showed a beneficial effect during the incubation period or earlier.

*PM*_2.5_ showed a significant impact around symptom onset, such that a higher *PM*_2.5_ of 11-18*μg*/*m*^3^ was associated with a higher OR of death (Figure 2C). The maximum OR was observed at symptom onset with a value of 1.72 (95% CI: 1.30-2.26) when *PM*_2.5_ reached 15*μg*/*m*^3^, compared with the reference (*PM*_2.5_ = 10*μg*/*m*^3^). The OR of these factors at specific infectious stages was described in the section *the effects of weather on the iCFR at specific time points* in Supplementary Information (see Figure S4).

### Cumulative and marginal effects of environmental factors

The cumulative effects of temperatures and *PM*_2.5_ were estimated for the duration between virus exposure and three days after symptom onset (or incubation period), whereas the cumulative effects of sunlight duration were estimated from three days prior to the virus exposure to three days after symptom onset (Figure 2D-F).

Overall, the cumulative effects (measured by log(OR)) of low temperatures (8-11°C) were higher than zero but not statistically significant (Figure 2D). Sunlight durations of 6-8h were significantly associated with a higher OR, while higher sunlight durations of >13h appear to be protective (Figure 2E). The cumulative effects of low *PM*_2.5_ (7-10 *μg*/*m*^3^) were significantly low and the effects of high *PM*_2.5_ (10-18 *μg*/*m*^3^) were substantially high (Figure 2F), suggesting a positive relationship between iCFR and *PM*_2.5_. While comparing the environmental predictors, the cumulative effect of the *PM*_2.5_ showed a larger variation than other predictors.

We further assessed the impacts on iCFR for one unit change in the predictor variables (Table 3). The iCFR increased by 102% for each *μg*/*m*^3^ rise in. *PM*_2.5_, whereas it decreased by 62% for one *μg*/*m*^3^ decrease. In contrast, the temperature and sunlight duration had an inverse effect on the risk of death, i.e., with one unit increase in temperature and sunlight duration reducing the risk of death by 8 and 69%, respectively. In comparison, a one-unit decline increases the risk by 51 and 98%.

**Table 3.**
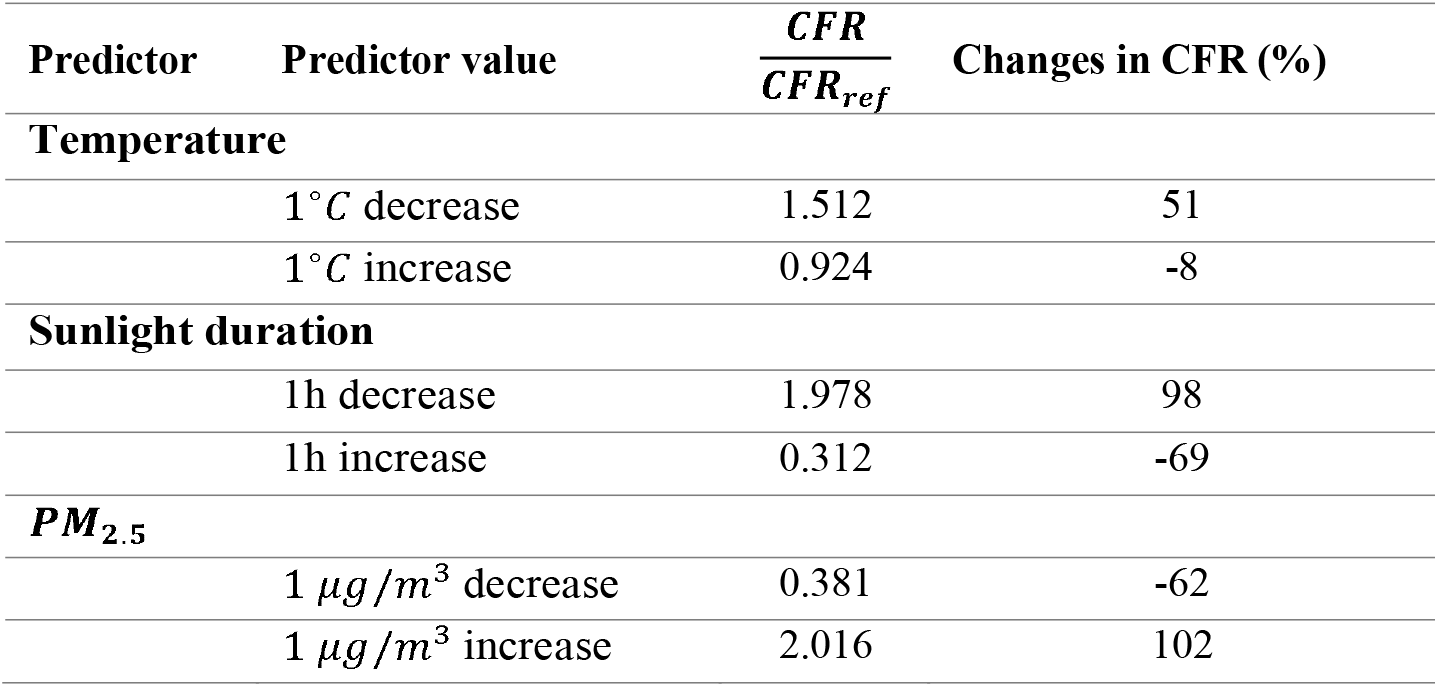
Changes in CFR under different scenarios of environmental predictors.

| Predictor | Predictor value | $\frac{CFR}{CFR_{ref}}$ | Changes in CFR (%) |
| --- | --- | --- | --- |
| **Temperature** |  |  |  |
|  | 1 °C decrease | 1.512 | 51 |
|  | 1 °C increase | 0.924 | -8 |
| **Sunlight duration** |  |  |  |
|  | 1h decrease | 1.978 | 98 |
|  | 1h increase | 0.312 | -69 |
| ***PM*<sub>2.5</sub>** |  |  |  |
| | 1 $\mu g/m^3$ decrease | 0.381 | -62 |
| | 1 $\mu g/m^3$ increase | 2.016 | 102 |

### Model validation

Finally, we validated Model 6 by predicting future iCFR between May 13, 2020 and September 15, 2020 (see Methods). The model was able to capture the trend among all countries. 81% of observed data were successfully predicted within 95% confidence interval (Figure 3).

**Figure 3.**
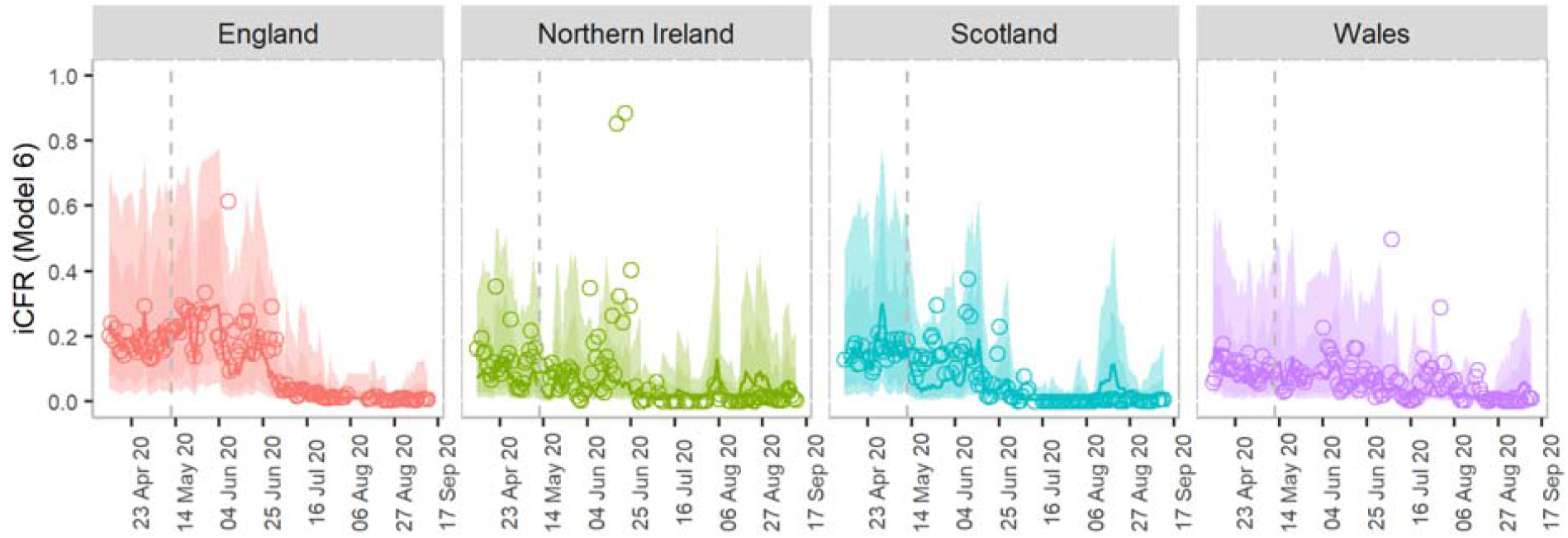
Sensitivity analysis of the best model (Model 6). The panels show the model’s prediction results until mid-September, the time when the first alpha variant has been detected in the UK. The points in each subplot represent the instantaneous CFR (iCFR) estimated using the back-projection method for each date. Solid lines represent the CFR estimated using the best-prediction model. The shaded regions indicate pointwise 75% and 95% prediction intervals, respectively. The vertical dashed lines represent the day until which we train the data in the model, whereas on the right side of the line are tested data.

## Discussion

Recent studies have shown that the mortality or CFR of COVID-19 was not only affected by the virulence of SARS-CoV-2 but also by environmental conditions, such as weather and air pollution (Benedetti et al. 2020; Liang et al. 2020; Ma et al. 2020; Wu et al. 2020b). However, whether their impacts were same across different infectious stages was unknown. After using the back-projection approach to obtain iCFR, we were able to provide evidence that lower temperature and sunlight exposure after symptom onset and increased *PM*_2.5_ around symptom onset resulted in a higher risk of death. This study employed a distributed lag nonlinear model, which enabled us to understand the lag effects of environmental variables to account for individual infectious statuses of infected cases (e.g., exposure period, incubation period, symptomatic period etc.). This finding suggests that different precautionary measures can be taken before and after symptom onset.

The results suggest that a specific range of temperature (e.g., between 8-°C) could increase the risk of COVID-19 death when patients were exposed to them after the incubation period. One possible reason is that exposure to cold temperature during these periods might deteriorate or influence COVID-19 patients’ immune responses (Liang and Yuan 2022; Schultze and Aschenbrenner 2021).

Sunlight appeared to have an important role in mortality. It affects the production of vitamin D (Haddad and Hahn 1973). Vitamin D deficiency results in impaired immune function, which can increase the risk of infectious diseases, such as those caused by respiratory viruses (Hart et al. 2011). Recent studies showed that there are no significant differences in hospital mortality between the vitamin D3 group and the placebo group (Leaf and Ginde 2021). A systemic review, however, investigated seven out of nine studies indicated that the lack of vitamin D greatly impacts the severity and death of COVID-19 (Yisak et al. 2021).

Prolonged exposure to sunlight has been found to inactivate SARS-CoV-2 (Chamary 2021; Ratnesar-Shumate et al. 2020), resulting in a reduced risk of infection or disease severity. However, whether prolonged exposure to sunlight may also suppress the proper functioning of the immune system is unknown, especially after the incubation period (Maglio et al. 2016). Our findings suggest the possible preventive effects of sunlight exposure on the disease severity of COVID-19 during but not after the incubation period.

In the UK, studies have revealed associations between the air pollutant *PM*_2.5_ and COVID-19 infection and mortality. For the existing COVID-19 cases, air pollutants, particularly *PM*_2.5_, may trigger airway inflammation. Ecological and individual-level investigation were conducted in the context of an association between air pollution and COVID-19 severity and mortality (Liang et al. 2020; Mendy et al. 2021; Wu et al. 2020a) and observed that the frequency of illness and fatalities rose significantly with the increment of the *PM*_2.5_ (Meo et al. 2021; Travaglio et al. 2021).

### Policy recommendations

During the past few years, after close contacts of an infected case are identified, many of them are quarantined until they are confirmed after testing, then immediately becoming home-isolated or hospital-isolated if they are COVID-19 positive cases. There is still no clear guideline on how to reduce disease severity after disease exposure besides clinical treatment. Certain recommendations can be given based on our findings.

1. For COVID-19 patients after symptoms appear, adopting certain preventive measures by maintaining the environment conditions (such as temperature, sunlight and *PM*_2.5_) in isolation facilities or at home may be effective in reducing the risk of severity.
2. Different preventive measures might need to be taken according to infectious stages. For example, after symptoms appear, infected individuals can maintain the environment with moderate or low sunlight. In contrast, during the incubation period, more sunlight shows a higher protective effect.

### Limitations

Recently, many studies have been conducted on the association between ambient temperatures and deaths where the inverse relationship was justified (Christophi et al. 2021; Liang and Yuan 2022; Wu et al. 2020b; Zhu et al. 2021). In contrast, at temperatures below 8°C the risk of death became low both before and after symptom onset. We were not able to exclude the possible influences of behavioral changes in the population during a very cold time. In cold temperatures, people usually stay more indoors and may be affected by the room temperatures rather than ambient temperatures. Although our study did not incorporate indoor temperature, a previous study suggested that indoor temperature was strongly correlated with outdoor temperature (Lee and Lee 2015). Therefore, the overall pattern of the risk of temperature might likely be similar even when indoor temperature is used.

Furthermore, the data used in this study were gathered during the initial lockdown period, which spanned from March to May 2020 without large variations in interventions. This period falls in the UK’s winter season, with colder temperatures and reduced sunlight predominating. Moreover, the study did not consider the population’s behavioral changes and indoor environments. For example, the use of heaters may influence the temperature in a room, etc. We suggest that further studies should be carried out to understand better the effects of environmental exposures on disease severity to capture all these limitations.

## Conclusions

For example, how to maintain proper environmental conditions, such as indoor temperature, sunlight, and air quality, during quarantine or home-isolation periods to reduce the probability of death is largely unknown. After many restrictions were lifted in many countries, people with COVID-19 symptoms are advised to get tested and self-isolate. Understanding the relationship between these environmental factors and iCFR especially after symptoms appear provides important suggestions for reducing the number of severe cases.

## Supporting information

COVID_19_death_UK_Main_20221022

## Data Availability

All data produced in the present work are contained in the manuscript.

## Acknowledgments

The authors are indebted to the City University of Hong Kong for providing excellent research facilities. The first author is also grateful to the university grant commission of Hong Kong for providing the most prestigious Hong Kong PhD Fellowship Scheme (HKPFS). Special thanks to Professor Antonio Gasparrini at the London School of Hygiene and Tropical Medicine and Professor Linwei Tian at the School of Public Health at the University of Hong Kong for the valuable feedback on the model development and manuscript formation.

## Statement and Declarations

### Funding

This work was supported by the grant number #9610416. Hsiang-Yu Yuan has received the support from the City University of Hong Kong.

### Competing Interest

The authors have no relevant financial or non-financial interests to disclosed.

## Author Contributions

All authors contributed to the study conception and design. The first draft of the manuscript was written by *M Pear Hossain* and all authors commented on previous versions of the manuscript. All authors read and approved the final manuscript.

*M. Pear Hossain*: Conceptualization; Data curation; Formal analysis; Methodology; Software; Validation; Visualization; Roles/Writing – original draft; Writing – review & editing.

*Wen Zhou*: Methodology; Writing – review & editing.

*Marco Y. T. Leung*: Writing – review & editing.

*Hsiang-Yu Yuan*: Conceptualization; Data curation; Funding acquisition; Supervision; Roles/Writing – original draft; Writing – review & editing.

## Data Availability

The data on COVID-19 cases and deaths and air pollution are publicly available in GOV.UK (https://www.gov.uk/coronavirus) and Air Information Resource in the Department for Environment Food and Rural Affairs, UK (https://uk-air.defra.gov.uk/) respectively. The weather data is also available in the public repository European Climate Assessment and Dataset (https://www.ecad.eu/).

## Ethics approval

The current study did not need any ethical approval since no animal model or organ was not used in this study.

## Consent to publish

All authors give their consent to publish this article.

## Notes

### Competing Interest Statement

The authors have declared no competing interest.

### Funding Statement

This work was supported by grant number #9610416. Hsiang-Yu Yuan has received support from the City University of Hong Kong.

