## Supplementary material for "Association between PM2.5 air pollution, temperature, and sunlight during different infectious stages with the case fatality of COVID-19 in the United Kingdom: a modeling study": COVID_19_death_UK_Main_20221022

**Name:** Hsiang-Yu Yuan

**Postal address:** 1A-311, 3/F, Block 1, To Yuen Building, 31 To Yuen Street, City University of Hong Kong, Hong Kong Special Administrative Region, China.

### Supplementary Information

###### S.1 Additional methods

The predictor-response relationship is expressed in equation (1) in the main text. The smooth functions of the weather predictors such as temperature and sunlight duration were presented by $s_{l}\left( X_{t,l}^{k}; \beta_{l} \right)$. Where $X_{t,l}^{k}$ represents the cross-basis matrix obtained by applying the distributed lag nonlinear model (DLNM) to weather (temperature and sunlight duration) and air pollution ($O_{3}$, $NO_{2}$, $SO_{2}$, $CO$, $PM_{10}$ and $PM_{2.5}$), $\beta_{l}$ is the vector of the coefficient of $X_{t,l}^{k}$,$l$ represents the lag days from the day of confirmation to the day of exposure and $L$ is the maximum lag, which was obtained by summing the incubation period and confirmation delay, i.e., $L=t_{1}+t_{2}$.

We used a natural cubic spline of each predictor variables to assess nonlinear effects, and an additional natural cubic spline of lag to evaluate the lag-response relationship. Spline knots for the weather predictors and lag were placed at equal distances within the predictors’ range and the lags’ log scale, respectively, to allow sufficient flexibility. Degrees of freedom (df) for both spline functions (predictors and lags) were chosen based on the Akaike information criterion (AIC). We examined a range of values between 1 and 10 with different combinations of spline dfs. We found that the best results were obtained for both weather predictors and lag at 4 and 5 df, respectively. All analyses were carried out using the data falling between the lower 2.5% and upper 97.5% of observations to avoid outlier bias.

###### S.2 Descriptive analysis

We conducted an exploratory data analysis and provided summary statistics for weather conditions, air pollution and COVID-19 data in the UK. We observed diverse weather conditions and air pollution across the UK during the first lockdown period. Scotland and Wales experienced the minimum and maximum temperatures (3.5 and 16.4^◦^C), respectively, while temperature variation (mean $\pm$ standard deviation (SD): $10.5 \pm2.7$) was greatest in England (Table S1). Wales had the most prolonged sunlight exposure, while Scotland experienced the least. In England and Wales, the median sunlight duration was almost 1.5-fold longer than that in Scotland. The variation in humidity was similar across all the UK countries.

England poses the maximum average concentration of $NO_{2}$ and fine particular matter $PM_{2.5}$, 14.7, and 12.3 $\mu g/m^{3}$ , respectively. Ozone concentration was observed maximum in Wales, whereas the difference in England was only four $\mu g/m^{3}$. On the other hand, Scotland is polluted mainly by SO2, while the CO concentration was equally distributed in all constituent countries.

In England, the CFR was almost double (0.22) of that in the other three countries (calculated as the ratio of deaths to cases in Table S1). The maximum number of daily number of cases exceeded 5,000 in England, while the minimum recorded number was 891 during the study period. Northern Ireland had the lowest daily incidence and number of deaths, with averages of 83 and 9 (CFR = 0.11), respectively. While the numbers of both cases and deaths were lowest in Northern Ireland, the lowest CFR was seen in Wales (0.10).

###### S.3 Variability in the instantaneous CFR and environmental predictors

We observed variation in both the iCFR and weather predictors for each country. The iCFR was highest at the beginning of the lockdown in each of the UK’s four countries (Figure S1). England had the highest risk of death, while Wales had the lowest. Interestingly, although iCFR values varied, the overall trend was similar in North Ireland, Scotland, and Wales.

From the beginning of the lockdown period, on March 26, temperatures trended upwards until mid-April (Figure S1). Temperatures fluctuated ± $1-2^{\circ}C$ after mid-April. Though similar trends were seen, temperatures varied across the geographical locations. Overall variation was the highest in England and Wales over the period studied.

Sunlight duration peaked during the second half of April in Scotland and Northern Ireland (Figure S1). From the beginning of the study period, sunlight duration trended upwards until the third week of April. However, a downward trend was observed in all four countries by the end of April. Though total sunlight duration varied, trends were similar across the countries studied. The largest variation was observed in Northern Ireland.

In contrast, the relative humidity followed an opposite trend to the other weather variables in the short term. Humidity trended downwards until the end of April, except for a small peak in early May. Humidity continued to decline from the following week until the end of the study period (Figure S1). The short-term trends of air pollution were also plotted (Figure S2).

###### S.4 Selecting weather and air pollution predictors and comparing models

We used a stepwise forward model selection approach to obtain the best-fitted model. Initially, we developed a baseline model incorporating just the regional random effects, which was adjusted for the days of the week. AIC values for the baseline model were very high (Table 1 in the main text). Then, we added cross-basis functions of each predictor variable individually to the baseline model using a distributed lag nonlinear model, which provided enough flexibility to explore the lag effects from the date of confirmation to the date of exposure. We considered three additional days for sunlight duration to investigate whether sunlight duration affects the iCFR beyond the incubation period.

We included variables in the model one after another. In the first step, we selected the model with sunlight duration as it provides the smallest AIC out of ten competitive models (Table S2). In addition to sunlight duration, we added the remaining nine variables into the model. The addition of more variables with cross-basis functions significantly reduced the AIC. This time model with sunlight duration and temperatures gives the best fitting model. We increased the complexity in the model if the AIC values improved. In the fourth step, we stopped including variables because the AIC value started rising after that.

To determine the best prediction model, we compared the three best-fitted candidate models. models. Given that all models compared in this study could explain the overall variation (i.e., the likelihood-ratio statistic equaled 1), the best-fitting models were chosen based on the AIC value. We found that Model 6 had the lowest AIC scores, while these scores were 1 unit higher for Model 5 and 8. Next, we compared Models 5, 6 and 8 based on the RMSE of the predicted results. The RMSE score was the smallest for Model 6 (Table 2 in the main text). Therefore, Model 6 was chosen as the best prediction model and was used for further analyses. This model successfully predicted the iCFR for different times and locations. Additionally, the model could provide a short-term trend in the iCFR in the UK. The prediction results for Models 5, 6 and 8 were depicted in the Figure S3.

###### S.5 The effects of weather on the iCFR at specific time points

We assessed the effects of temperature, sunlight duration, and $PM_{2.5}$ at specific points in the course of disease infection, particularly near virus exposure, during symptom onset, and near case confirmation (two days later the symptom onset). The temperatures were found to significantly affect the iCFR two days after symptom onset with values of 9-${11}^{\circ}C$ (Figure S4). Lower exposure to sunlight duration, such as 5-10h, showed high OR near virus exposure and symptom onset. In contrast, higher $PM_{2.5}$ at and after symptom onset was more likely to increase the risk of deaths.

###### S.6 List of supplementary figures

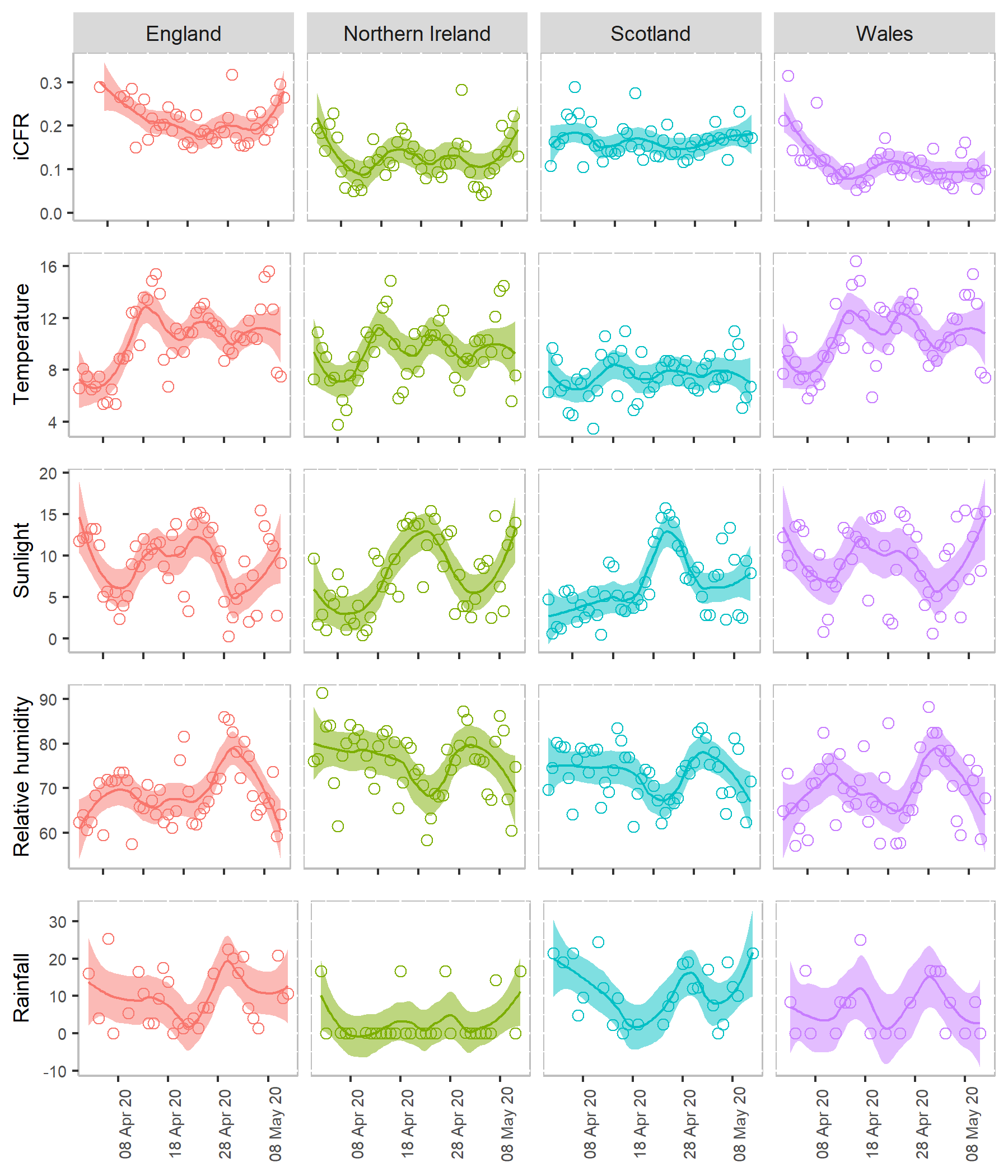

Figure S1. **Time-varying instantaneous case fatality rates (iCFRs) and weather factors (temperature, sunlight, relative humidity, and rainfall) between 26 March and 12 May 2020 in the UK**. iCFR is defined as a ratio of the retrospectively adjusted deaths and reported cases. Temperature was measured in °$C$, and sunlight in hours, humidity in %, and rainfall in $mm$ . A LOESS smoothing function was used to obtain a smooth line representing the trend over time. The shaded region indicates the pointwise 95% confidence interval. Circles represent the estimated CFR and weather factors observed on each day.

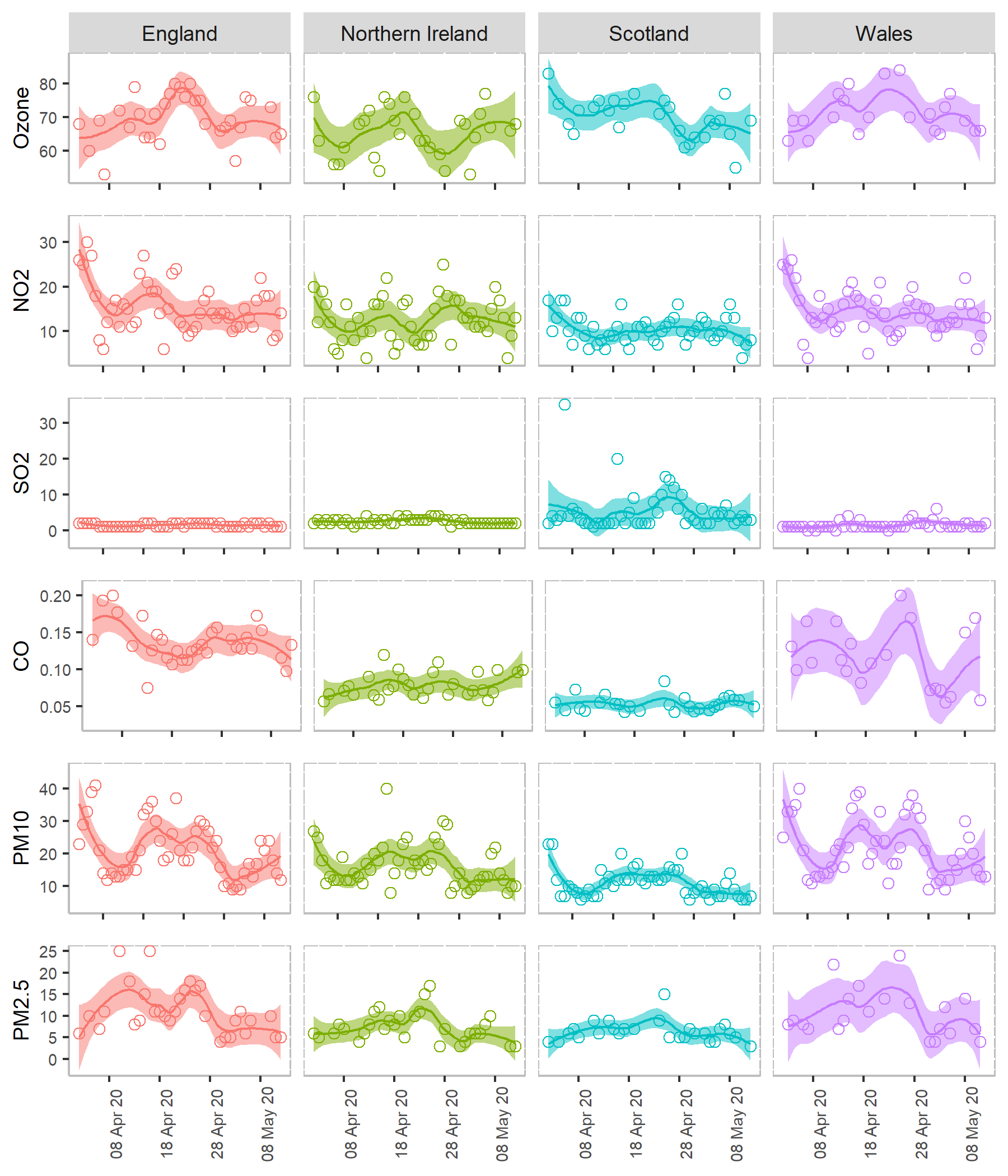

Figure S2. **Air pollutants (ozone,** $\boldsymbol{N}\boldsymbol{O}_{\boldsymbol{2}}$***,*** $\boldsymbol{S}\boldsymbol{O}_{\boldsymbol{2}}$***,*** $\boldsymbol{CO}$***,*** $\boldsymbol{P}\boldsymbol{M}_{\boldsymbol{10}}$ **and** $\boldsymbol{P}\boldsymbol{M}_{\boldsymbol{2.5}}$ **between 26 March and 12 May 2020. Each air pollutant was measured in** $\boldsymbol{\mu g/}\boldsymbol{m}^{\boldsymbol{3}}$. A LOESS smoothing function was used to obtain a smooth line representing the trend over time. The shaded regions indicate the pointwise 95% confidence interval. Circles represent the estimated CFR, weather factors and air pollutants observed on each day.

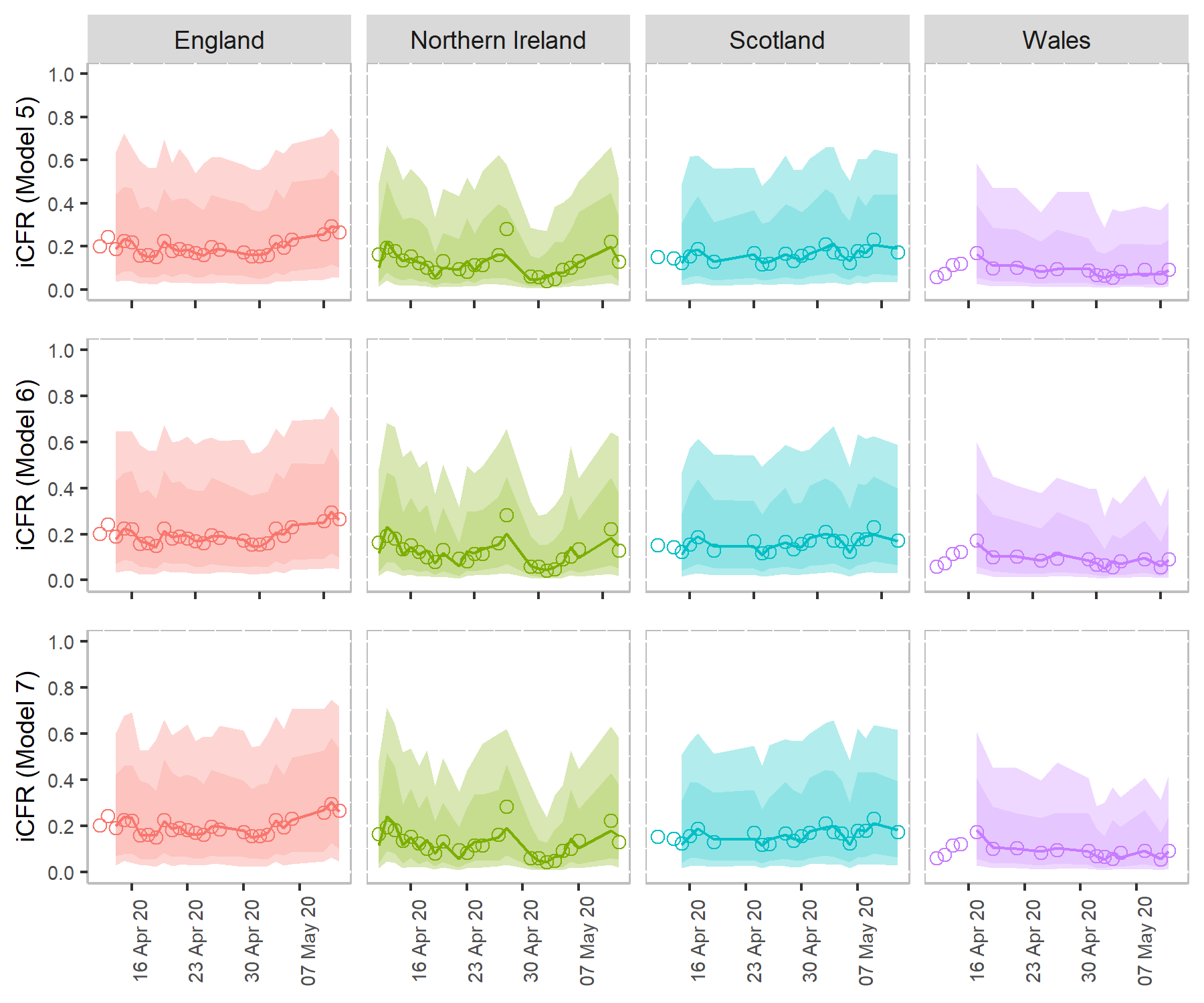

Figure S3. **Comparing the predicted results of instantaneous case fatality rates in the UK using three candidate models (Model 5, 6 and 7).** Model 6 referred as the best model. The points in each subplot represent the retrospectively adjusted iCFR for each date. Lines represent the CFR estimated using the candidate models, as provided in Table 2 in the main text. The shaded regions indicate pointwise 75% and 95% prediction intervals respectively.

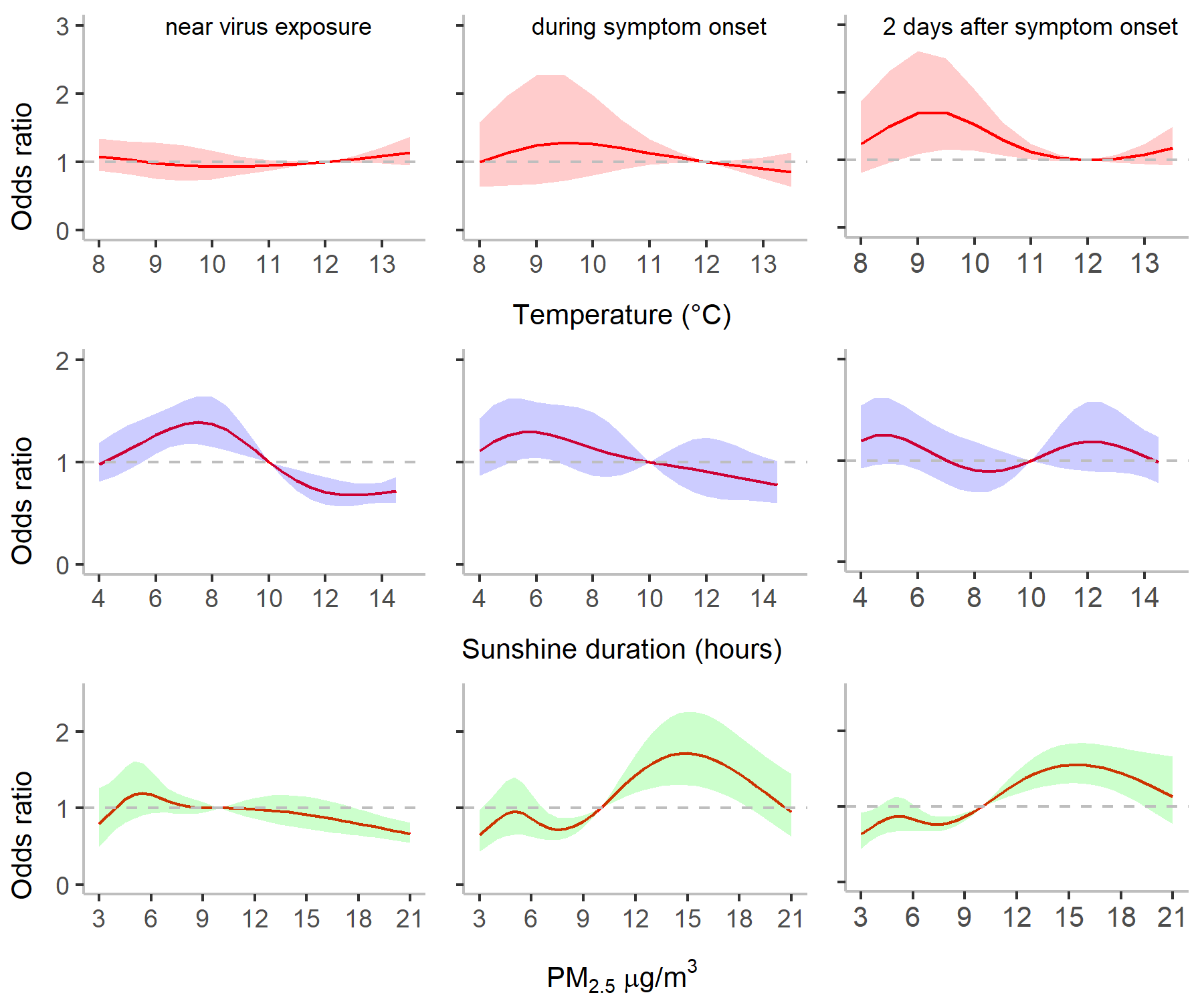

Figure S4. **Effects of temperature, sunlight duration and** $\boldsymbol{P}\boldsymbol{M}_{\boldsymbol{2.5}}$**on instantenious case fatality rate at specific time points.** The column ‘near virus exposure’ represents day 0, ‘during symptom onset’ represents day 6, and ‘2 days after symptom onset’ represents day 8 since virus exposure. Horizontal lines represent the baseline odds ratio at the reference values of the weather conditions and air pollution. We considered the time points closest to case confirmation, symptom onset and exposure.

###### S.7 List of supplementary tables

Table S1. **Summary statistics of COVID-19 infection and environmental risk factors in the UK.** Mean and standard deviation (SD), and five summary measures, such as minimum (Min), maximum (Max), median and 25^th^ ($P_{25}$) and 75^th^ ($P_{75}$) percentiles, for each variable are presented.

| Type | Variables | Country | Mean | SD | Min | $P_{25}$ | Median | $P_{75}$ | Max |
| --- | --- | --- | --- | --- | --- | --- | --- | --- | --- |
| COVID-19 | Daily cases | England | 2712 | 981 | 891 | 2050 | 2666 | 3555 | 5107 |
|  |  | Northern Ireland | 83 | 35 | 30 | 53 | 84 | 108 | 159 |
|  |  | Scotland | 272 | 85 | 136 | 185 | 281 | 338 | 430 |
|  |  | Wales | 228 | 95 | 16 | 158 | 233 | 291 | 502 |
|  | Daily deaths | England | 597 | 261 | 169 | 364 | 605 | 742 | 1123 |
|  |  | Northern Ireland | 9 | 6 | 1 | 4 | 7 | 13 | 30 |
|  |  | Scotland | 38 | 28 | 1 | 10 | 42 | 59 | 84 |
|  |  | Wales | 23 | 19 | 4 | 13 | 18 | 28 | 110 |
| Weather | Temperatures | England | 10.5 | 2.7 | 5.4 | 8.9 | 10.6 | 12.4 | 15.6 |
|  |  | Northern Ireland | 9.4 | 2.5 | 3.8 | 7.6 | 9.4 | 10.7 | 14.9 |
|  |  | Scotland | 7.5 | 1.7 | 3.5 | 6.4 | 7.4 | 8.7 | 11.0 |
|  |  | Wales | 10.7 | 2.6 | 5.8 | 8.7 | 10.3 | 12.7 | 16.4 |
|  | Sunlight | England | 8.9 | 4.2 | 0.3 | 5.1 | 9.6 | 12.2 | 15.5 |
|  |  | Northern Ireland | 7.8 | 4.6 | 0.4 | 3.9 | 7.8 | 12.2 | 15.4 |
|  |  | Scotland | 6.8 | 4.0 | 0.5 | 3.5 | 5.7 | 9.3 | 15.8 |
|  |  | Wales | 9.4 | 4.5 | 0.6 | 6.4 | 10.1 | 13.3 | 15.5 |
|  | Humidity | England | 69.4 | 6.7 | 57.5 | 64.2 | 68.7 | 72.3 | 86.0 |
|  |  | Northern Ireland | 75.6 | 7.2 | 58.4 | 70.5 | 76.5 | 80.4 | 87.3 |
|  |  | Scotland | 73.2 | 6.1 | 61.3 | 68.6 | 73.7 | 78.3 | 83.6 |
|  |  | Wales | 70.0 | 8.1 | 57.1 | 64.6 | 70.1 | 76.1 | 88.2 |
|  | Rainfall | England | 10.8 | 7.8 | 0.0 | 3.7 | 9.5 | 16.7 | 25.4 |
|  |  | Northern Ireland | 4.8 | 8.2 | 0.0 | 0.0 | 0.0 | 14.3 | 33.3 |
|  |  | Scotland | 11.6 | 7.8 | 0.0 | 4.2 | 12.2 | 18.6 | 24.4 |
|  |  | Wales | 8.2 | 8.9 | 0.0 | 0.0 | 8.3 | 16.7 | 33.3 |
| Air pollutants | O3 | England | 69.6 | 7.2 | 53.0 | 64.0 | 68.5 | 75.3 | 83.0 |
|  |  | Northern Ireland | 65.1 | 9.2 | 37.0 | 59.8 | 66.5 | 71.3 | 82.0 |
|  |  | Scotland | 70.6 | 6.8 | 50.0 | 67.0 | 71.0 | 75.3 | 83.0 |
|  |  | Wales | 74.2 | 9.3 | 55.0 | 68.8 | 73.0 | 81.3 | 97.0 |
|  | NO2 | England | 14.7 | 5.1 | 6.0 | 11.0 | 14.0 | 18.0 | 27.0 |
|  |  | Northern Ireland | 12.3 | 4.9 | 4.0 | 8.8 | 12.0 | 16.0 | 25.0 |
|  |  | Scotland | 10.2 | 3.0 | 4.0 | 8.0 | 10.0 | 12.0 | 17.0 |
|  |  | Wales | 13.5 | 4.3 | 4.0 | 11.0 | 13.0 | 16.3 | 22.0 |
|  | SO2 | England | 1.4 | 0.5 | 1.0 | 1.0 | 1.0 | 2.0 | 2.0 |
|  |  | Northern Ireland | 2.6 | 0.8 | 1.0 | 2.0 | 2.0 | 3.0 | 4.0 |
|  |  | Scotland | 5.4 | 5.9 | 1.0 | 2.0 | 4.0 | 6.0 | 35.0 |
|  |  | Wales | 1.5 | 1.1 | 0.0 | 1.0 | 1.0 | 2.0 | 6.0 |
|  | CO | England | 0.1 | 0.0 | 0.1 | 0.1 | 0.1 | 0.2 | 0.2 |
|  |  | Northern Ireland | 0.1 | 0.0 | 0.0 | 0.1 | 0.1 | 0.1 | 0.1 |
|  |  | Scotland | 0.1 | 0.0 | 0.0 | 0.0 | 0.1 | 0.1 | 0.1 |
|  |  | Wales | 0.1 | 0.1 | 0.1 | 0.1 | 0.1 | 0.2 | 0.3 |
|  | PM10 | England | 20.5 | 8.3 | 9.0 | 14.0 | 18.5 | 24.5 | 41.0 |
|  |  | Northern Ireland | 15.8 | 6.5 | 8.0 | 11.8 | 14.0 | 18.3 | 40.0 |
|  |  | Scotland | 10.5 | 3.7 | 6.0 | 7.0 | 10.0 | 13.0 | 20.0 |
|  |  | Wales | 21.3 | 9.2 | 9.0 | 13.8 | 20.0 | 29.3 | 40.0 |
|  | PM2.5 | England | 12.3 | 6.7 | 4.0 | 7.0 | 10.5 | 16.0 | 30.0 |
|  |  | Northern Ireland | 7.0 | 3.8 | 2.0 | 4.8 | 6.0 | 8.0 | 22.0 |
|  |  | Scotland | 6.0 | 2.6 | 3.0 | 4.0 | 6.0 | 7.0 | 15.0 |
|  |  | Wales | 12.0 | 7.3 | 3.0 | 6.8 | 9.5 | 15.5 | 29.0 |

Table S2. **Stepwise forward model selection.** $T, S, R$ and $H$ represent the weather predictors temperatures, sunlight duration, rainfall, and humidity. $f(.)$ represents cross-basis function of each predictor variables. At each step, bold fonts represent a best candidate model. The model Tri7 and Tri8 at the step 3 and Tetra2 at the step 4 were selected as three best-fitting models with low AIC.

| Step | Model No. | Variables | AIC | Significant variables |
| --- | --- | --- | --- | --- |
| 1 | U1 | $f(T)$ | 772 | $T$ |
|  | **U2** | $\boldsymbol{f(S)}$ | **650** | $\boldsymbol{S}$ |
|  | U3 | $f(R)$ | 751 | $R$ |
|  | U4 | $H_{t-(t_{1}+t_{2})}$ | 1149 | - |
|  | U5 | $f\left( O_{3} \right)$ | 779 | $O_{3}$ |
|  | U6 | $f(NO_{2})$ | 753 | $NO_{2}$ |
|  | U7 | $f(SO_{2})$ | 832 | - |
|  | U8 | $f\left( CO \right)$ | 765 | $CO$ |
|  | U9 | $f(PM_{10})$ | 695 | $PM_{10}$ |
|  | U10 | $f\left( PM_{2.5} \right)$ | 736 | $PM_{2.5}$ |
| 2 | **Bi1** | $\boldsymbol{f}\left( \boldsymbol{S} \right)\boldsymbol{+f(T)}$ | **595** | $\boldsymbol{T+S}$ |
|  | Bi2 | $f\left( S \right)+f(R)$ | 613 | $S+R$ |
|  | Bi3 | $f\left( S \right)+H_{t-(t_{1}+t_{2})}$ | 651 | $S$ |
|  | Bi4 | $f\left( S \right)+f(O_{3})$ | 596 | $S+O_{3}$ |
|  | Bi5 | $f\left( S \right)+f(NO_{2})$ | 600 | $S+NO_{2}$ |
|  | Bi6 | $f\left( S \right)+f(SO_{2})$ | 620 | $S+SO_{2}$ |
|  | Bi7 | $f\left( S \right)+f(CO)$ | 645 | $S+CO$ |
|  | Bi8 | $f\left( S \right)+f(PM_{10})$ | 608 | $S+PM_{10}$ |
|  | Bi9 | $f\left( S \right)+f(PM2.5)$ | 616 | $S+PM_{2.5}$ |
| 3 | Tri1 | $f\left( S \right)+f\left( T \right)+f(R)$ | 590 | $S$ |
|  | Tri2 | $f\left( S \right)+f\left( T \right)+H_{t-\left( t_{1}+t_{2} \right)}$ | 596 | $T+S$ |
|  | Tri3 | $f\left( S \right)+f\left( T \right)+f(O_{3})$ | 589 | $T+S$ |
|  | Tri4 | $f\left( S \right)+f\left( T \right)+f(NO_{2})$ | 584 | $S+NO_{2}$ |
|  | Tri5 | $f\left( S \right)+f\left( T \right)+f\left( SO_{2} \right)$ | 581 | $T+S+SO_{2}$ |
|  | Tri6 | $f\left( S \right)+f\left( T \right)+f(CO)$ | 590 | $T+S$ |
|  | Tri7 | $f\left( S \right)+f\left( T \right)+f(PM_{10})$ | 579 | $T+S+PM_{10}$ |
|  | **Tri8** | $\boldsymbol{f}\left( \boldsymbol{S} \right)\boldsymbol{+f}\left( \boldsymbol{T} \right)\boldsymbol{+f(P}\boldsymbol{M}_{\boldsymbol{2.5}}\boldsymbol{)}$ | **578** | $\boldsymbol{T+S+P}\boldsymbol{M}_{\boldsymbol{2.5}}$ |
| 4 | Tetra1 | $f\left( S \right)+f\left( T \right)+f\left( PM_{2.5} \right)+f(R)$ | 594 | $T$ |
|  | **Tetra2** | $\boldsymbol{f}\left( \boldsymbol{S} \right)\boldsymbol{+f}\left( \boldsymbol{T} \right)\boldsymbol{+f}\left( \boldsymbol{P}\boldsymbol{M}_{\boldsymbol{2.5}} \right)\boldsymbol{+}\boldsymbol{H}_{\boldsymbol{t-(}\boldsymbol{t}_{\boldsymbol{1}}\boldsymbol{+}\boldsymbol{t}_{\boldsymbol{2}}\boldsymbol{)}}$ | **579** | $\boldsymbol{T+S+P}\boldsymbol{M}_{\boldsymbol{2.5}}$ |
|  | Tetra3 | $f\left( S \right)+f\left( T \right)+f\left( PM_{2.5} \right)+f(O_{3})$ | 592 | $T+S$ |
|  | Tetra4 | $f\left( S \right)+f\left( T \right)+f\left( PM_{2.5} \right)+f\left( NO_{2} \right)$ | 592 | $S+PM_{2.5}$ |
|  | Tetra5 | $f\left( S \right)+f\left( T \right)+f\left( PM_{2.5} \right)+f\left( SO_{2} \right)$ | 587 | $T+S+PM_{2.5}$ |
|  | Tetra6 | $f\left( S \right)+f\left( T \right)+f\left( PM_{2.5} \right)+f\left( CO \right)$ | 594 | $T+S$ |
|  | Tetra7 | $f\left( S \right)+f\left( T \right)+f\left( PM_{2.5} \right)+f(PM_{10})$ | 595 | - |
